# Evaluating Large Language Models for Transparent Quality-of-Care Measurement in Children with ADHD

**DOI:** 10.64898/2026.04.12.26350732

**Authors:** Yair Bannett, Malvika Pillai, Tracy Huang, Ingrid Luo, Fatma Gunturkun, Tina Hernandez-Boussard

**Affiliations:** Division of Developmental-Behavioral Pediatrics, Stanford University School of Medicine, Stanford, California, USA; Biomedical Informatics Research Center, Stanford University School of Medicine, Stanford, California, USA; Quantitative Sciences Unit, Stanford University School of Medicine, Stanford, CA, USA

**Keywords:** Artificial intelligence, Large language models, Quality-of-Care, Attention-Deficit/Hyperactivity Disorder, Pediatrics

## Abstract

**Importance:** Guideline-concordant care for young children with attention-deficit/hyperactivity disorder (ADHD) includes recommending parent training in behavior management (PTBM) as first-line treatment. However, assessing guideline adherence through manual chart review is time-consuming and costly, limiting scalable and timely quality-of-care measurement.

**Objective:** To evaluate the accuracy and explainability of large language models (LLMs) in identifying PTBM recommendations in pediatric electronic health record (EHR) notes as a scalable alternative to manual chart review.

**Design, Setting, and Participants:** This retrospective cohort study was conducted in a community-based pediatric healthcare network in California consisting of 27 primary care clinics. The study cohort included children aged 4-6 years with ≥ 2 primary care visits between 2020-2024 and ICD-10 diagnoses of ADHD or ADHD symptoms (n=542 patients). Clinical notes from the first ADHD-related visit were included. A stratified subset of 122 notes, including all cases with model disagreement, was manually annotated to assess model performance in identifying PTBM recommendations and rank model explanations.

**Exposures:** Assessment and plan sections of clinical notes were analyzed using three generative large language models (Claude-3.5, GPT-4o, and LLaMA-3.3-70B) to identify the presence of PTBM recommendations and generate explanatory rationales and documentation evidence.

**Main Outcomes and Measures:** Model performance in identifying PTBM recommendations (measured by sensitivity, positive predictive value (PPV), and F1-score) and qualitative explainability ratings of model-generated rationales (based on the QUEST framework).

**Results:** All three models demonstrated high performance compared to expert chart review. Claude-3.5 showed balanced performance (sensitivity=0.89, PPV=0.95, and F1-score=0.92) and ranked highest in explainability. LLaMA3.3-70B achieved sensitivity=0.91, PPV=0.89, and F1-score=0.90, ranking second for explainability. GPT-4o had the highest PPV [0.97] but lowest sensitivity [0.82], with an F1-score of 0.89 and the lowest explainability ranking. Based on classifications from the best-performing model, Claude-3.5, 26.4% (143/542) of patients had documented PTBM recommendations at their first ADHD-related visit.

**Conclusions and Relevance:** LLMs can accurately extract guideline-concordant clinician recommendations for non-pharmacological ADHD treatment from unstructured clinical notes while providing clear explanations and supporting evidence. Evaluating model explainability as part of LLM implementation for medical chart review tasks can promote transparent and scalable solutions for quality-of-care measurement.

## INTRODUCTION

Attention-deficit/hyperactivity disorder (ADHD) is the most common neurobehavioral disorder in childhood, affecting an estimated 8-10% of U.S. children.^1^ Primary care pediatricians (PCP) play a central role in ADHD management, particularly in young children.^2,3^ For preschool-aged children (4-5 years), the American Academy of Pediatrics clinical practice guidelines recommend parent training in behavior management (PTBM) as first-line treatment because of stronger evidence for the effectiveness of behavioral therapy than pharmacotherapy in this age group.^4,5^ Despite this guidance, recent studies have found low rates of PTBM recommendations documented by PCPs, as well as evidence of premature use of psychoactive medications for preschoolers with ADHD^6–8^ – a practice pattern that may contribute to adverse short- and long-term outcomes for children and families.^9^

Learning health systems require efficient and scalable methods to monitor the quality of clinical care in order to address gaps in care, mitigate patient harm, and improve patient outcomes. However, there is currently no such method to assess guideline-concordant care for ADHD and other child mental health conditions. Health organizations rely either on claims data, which capture limited aspects of care, or on manual chart review of electronic health records (EHRs), which is costly, labor-intensive, and not scalable.^10–12^ Although artificial intelligence (AI) tools using natural language processing (NLP) of clinical text have been proposed to support quality-of-care assessment,^13–16^ adoption has been limited due to concerns about model transparency, interpretability, and trustworthiness.^17^ Additionally, traditional NLP models provide “black box” outputs without explanations, limiting their utility for clinician feedback and quality improvement.^18^

Recent advances in reasoning capabilities of generative large language models (LLMs) present a unique opportunity to offer transparent and scalable quality assessment. Unlike traditional NLP models, LLMs can be prompted to classify free-text notes and generate human-readable rationales for their classifications. Leveraging these capabilities to develop and test AI tools with clinician-facing explanations may overcome adoption barriers and improve trust and acceptability of AI-powered quality-of-care measurement.^19^

In this study, we evaluated the use of generative LLMs (specifically Claude-3.5, GPT-4o, and LLaMA3.3-70B) as a scalable and interpretable approach for identifying PTBM recommendations in pediatric ADHD care. We combined quantitative evaluation of model classification performance with qualitative evaluation of a model’s ability to provide clinically interpretable explanations for their classifications, using a comprehensive framework for human evaluation of LLMs.^20^ This work establishes the feasibility of leveraging LLMs for scalable, transparent quality-of-care measurement, providing actionable feedback for clinicians to enhance evidence-based behavioral health care for children.

## METHODS

### Study Setting and Population

This study was conducted within Packard Children’s Health Alliance (PCHA), a community-based pediatric network in the San Francisco Bay Area affiliated with Stanford Medicine Children’s Health and Lucile Packard Children’s Hospital. The network comprises 27 primary-care pediatric clinics. All visit diagnoses are clinician-entered across sites, ensuring reliability of cohort identification via diagnostic codes.^21^ This study was approved by the Stanford University School of Medicine Institutional Review Board. The full study process is illustrated in **Figure 1**.

**Figure 1.**
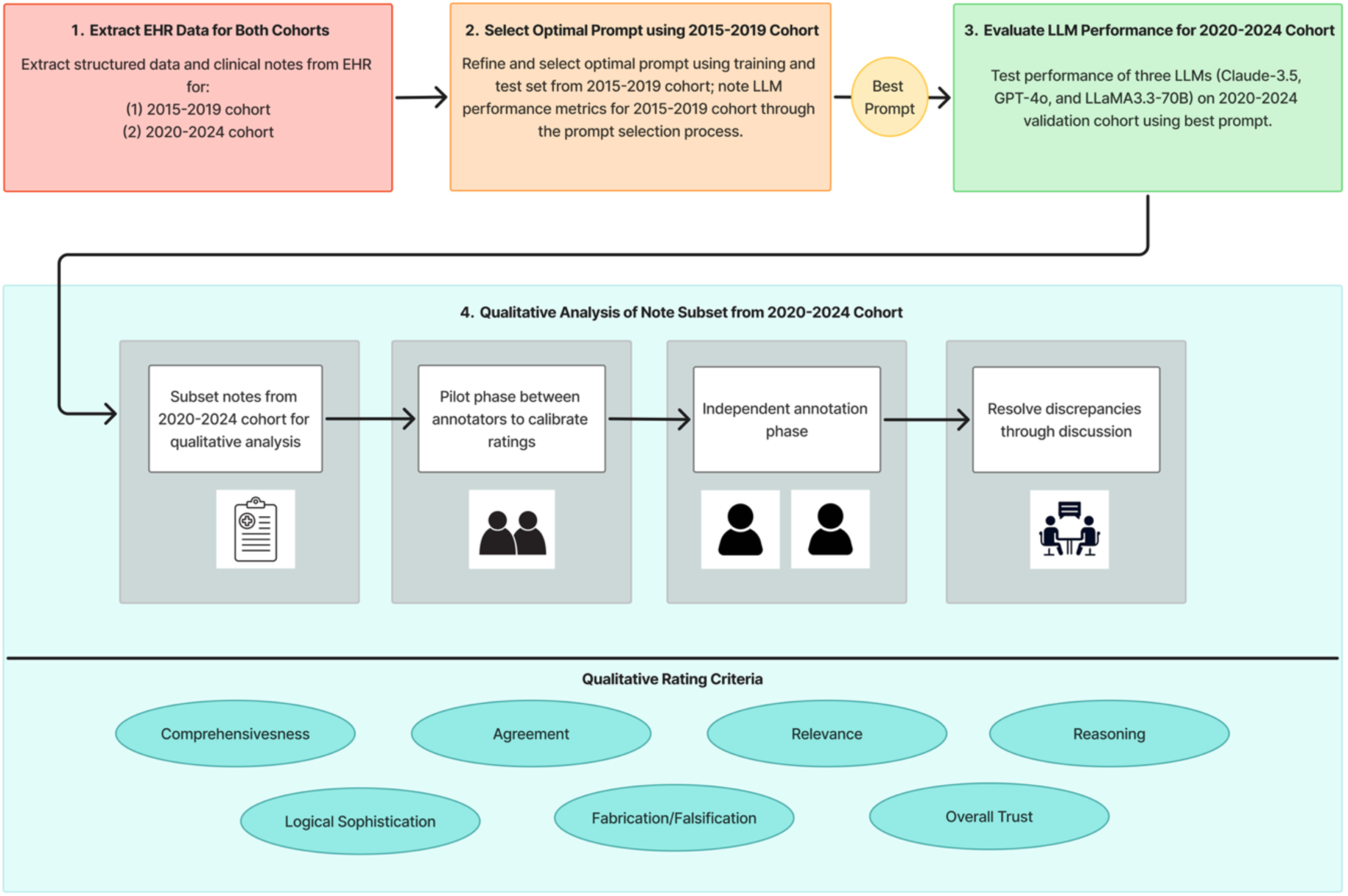
Study Flowchart Process

### Study Design, Data Sources, and Cohort Selection

We retrospectively extracted EHR free-text clinical notes for children aged 4 to 6 years with at least two primary care visits and an ICD-10 diagnosis of ADHD or ADHD-related symptoms (**Supplementary eTable 1**). The study included two temporally distinct cohorts: (1) Historical cohort (2015-2019) comprising 423 children, described in our prior study,^15^ and (2) Validation cohort (2020-2024), comprising 542 children, selected using the same inclusion criteria. Patients from the historical cohort were excluded from the validation cohort. We also excluded patients with an autism diagnosis in both cohorts, as evidence-based treatment for children with autism is not PTBM.

### Clinical Note Data Preparation

Clinical notes were organized in a Subjective/Objective/Assessment/Plan (SOAP) structure. We focused on the “Assessment and Plan” sections because they contain clinicians’ impressions and treatment recommendations. For each patient, we extracted the “Assessment and Plan” section from the progress note of the first ADHD-related visit (i.e., visit with an ADHD diagnosis) using an LLM-based sectionizer with a zero-shot prompt (LLaMA3.3-70B). In cases where the sectionizer produced errors, notes were processed using a rule-based sectionizer and manually reviewed.

### Prompt Engineering and Model Deployment

We evaluated three large language models: Claude-3.5 [Anthropic], GPT-4o [OpenAI], and LLaMA3.3-70B [Meta]. All models were accessed via a secure environment during a fixed evaluation period (May 2025).

We developed and tested three prompt variants: (1) basic zero-shot prompt asking whether the note included a PTBM recommendation or not; (2) detailed zero-shot prompt incorporating explicit PTBM definitions and inclusion/exclusion criteria; and (3) chain-of-thought prompt instructing the model to explain its reasoning prior to classification. All prompts required models to provide a binary classification of whether clinical notes contained a PTBM recommendation and generate explanations for their classifications linked to direct evidence from the note. PTBM recommendations were defined as referral to a therapist who provides PTBM and/or counseling the family in the office regarding behavior management principles.

Prompts were refined using the historical cohort training set (n=296), described in our prior study.^15^ To identify the best-performing prompt approach across models, a final version of each of the three prompt variants was evaluated on a separate, previously annotated, hold-out test set from the historical cohort (n=127 notes). The detailed zero-shot prompt demonstrated the highest F1 score across models and was selected for subsequent analyses. Using the same prompt (**Supplementary eTable 2**), the three models were prompted to classify notes from the historical cohort test set (n=127) and all notes from the validation cohort (n=542).

### Sampling of Notes from the Validation Cohort for Manual Annotation

To sample notes for manual annotation, we leveraged available model classifications and included all notes with model classification disagreement (n=38), aiming to manually review potentially ambiguous cases. For notes with unanimous classifications, sample sizes were calculated separately for the positive and negative groups using class-specific performance estimates from the historical test set. For the positive group, the calculation was based on the lowest true positive rate across models (0.80), and for the negative group, it was based on the lowest true negative rate (0.94). To achieve a ±10% margin of error at 95% confidence, we sampled 62 unanimously positive notes and 22 unanimously negative notes, resulting in a total of 122 notes in the test set.

### Manual Annotation of Clinical Notes in Validation Cohort Test Set

As done in our prior study,^15^ two trained annotators (pediatrician and trained researcher) independently annotated extracted sections of clinical notes (n=122) to determine whether the note contained mention of PTBM recommendations using pre-defined annotation guidelines. After establishing high inter-annotator agreement on a random sample of 20 notes (F-measure=0.90), the remaining notes were divided and annotated independently. All notes with model disagreement were double annotated and discrepancies were resolved through consensus, with a second pediatrician adjudicating unresolved cases.

### Model Quantitative Evaluation and Temporal Validation

Model performance was evaluated against manually annotated ground-truth labels in both the historical cohort test set (n=127 notes) and the validation cohort test set (n=122 notes), which assessed temporal generalizability. Performance metrics included sensitivity, positive predictive value (PPV), and F1-score. Descriptive statistics and a two-proportion z-test were used to summarize and compare the estimated rate of PTBM recommendations in the full historical (n=423) and validation (n=542) cohorts.

### Qualitative Evaluation of LLM-Generated Rationales in the Validation Cohort (2020-2024)

Guided by the QUEST framework,^20^ we evaluated the extent to which LLM-generated explanations (i.e., the rationale for its PTBM classification) provide reliable information. The QUEST framework includes five domains: Quality of information, Understanding and reasoning, Expression style and persona, Safety and harm, and Trust and confidence. Within these domains, we selected six criteria most relevant to clinical interpretability in the examined task: (1) comprehensiveness (addressed all relevant aspects of classification task), (2) relevance (did not include unrelated or unnecessary information), (3) agreement (no contradictions within the explanation, or between explanation and classification), (4) reasoning (provided clear logic linking evidence to classification), (5) fabrication/falsification (no incorrect or unsupported statements), and (6) overall trust (explanation promoted annotator trust in model classification). A supplemental criterion outside the QUEST framework was included to capture logical sophistication (synthesized and interpreted evidence in balanced, context-aware manner). Detailed annotation guidelines for qualitative rating were created for each criterion with modified instructions for false positive, true positive, false negative, and true negative cases (**Supplementary eTable 3**).

### Sampling and Ranking of Model Explanations

We purposefully sampled 54 LLM-generated explanations from the validation cohort, including 10 unanimously true-negative notes, 10 unanimously true-positive notes, and 34 notes in which at least one LLM disagreed with ground truth labels. After joint annotation of 5 cases, the two annotators independently rated the remaining cases using binary or three-point scales to assess comprehensiveness, relevance, agreement, reasoning, logical sophistication, fabrication/falsification, and overall trust (**Supplementary eTable 3**). Discrepancies were resolved through consensus meetings.

All ratings were subsequently normalized on a 0-1 scale, with higher values indicating better performance. The mean normalized score for each criterion was calculated separately for positive and negative prediction categories. For overall trust, the mean score was calculated across all cases, irrespective of prediction category.

## RESULTS

### Cohort Characteristics

The historical cohort (2015-2019) included 423 children aged 4 to 6 years with a diagnosis of ADHD (disorder or symptom-level) and at least two primary care visits. Applying the same inclusion criteria and excluding patients from the historical cohort, the validation cohort (2020-2024) included 542 children. **Table 1** presents the sociodemographic characteristics of the validation cohort. Of 542 patients, 69.9% (n=379) were male, 38.2% (n=207) were non-Hispanic White, and 77.5% (n=420) were privately insured.

**Table 1.** Characteristics of 2020-2024 Patient Cohort.

| Characteristics | Overall<br>(n=542) | PTBM Recommended<br>(n=143) | PTBM Not Recommended<br>(n=399) |
| --- | --- | --- | --- |
| Sex, N (%) |  |  |  |
| Male | 379 (69.9%) | 107 (74.8%) | 272 (68.2%) |
| Female | 163 (30.1%) | 36 (25.2%) | 127 (31.8%) |
| Race, N (%) |  |  |  |
| Non-Hispanic White | 207 (38.2%) | 57 (39.9%) | 150 (37.6%) |
| Non-Hispanic Asian | 107 (19.7%) | 29 (20.3%) | 78 (19.5%) |
| Non-Hispanic Black | 27 (5.0%) | 8 (5.6%) | 19 (4.8%) |
| American Indian or<br>Alaska Native | 3 (0.6%) | 2 (1.4%) | 1 (0.3%) |
| Unknown | 198 (36.5%) | 47 (32.9%) | 151 (37.8%) |
| Ethnicity, N (%) |  |  |  |
| Hispanic | 77 (14.2%) | 18 (12.6%) | 59 (14.8%) |
| Non-Hispanic | 366 (67.5%) | 101 (70.6%) | 265 (66.4%) |
| Unknown | 99 (18.3%) | 24 (16.8%) | 75 (18.8%) |
| Insurance, N (%) |  |  |  |
| Private | 420 (77.5%) | 109 (76.2%) | 311 (77.9%) |
| Public | 121 (22.3%) | 34 (23.8%) | 87 (21.8%) |
| Unknown | 1 (0.2%) | 0 (0.0%) | 1 (0.3%) |

PTBM = Parent training in behavior management

### Rate of Documented PTBM Recommendations

In the validation cohort, based on classifications from the best-performing model, Claude-3.5, 26.4% (143/542) of patients received a documented recommendation for PTBM at their first ADHD-related visit, which was not statistically significantly different from the rate found in the historical cohort (30.5%, 129/423; p=0.16).

### LLM Quantitative Evaluation: Historical Cohort (2015-2019)

Using the same selected prompt, all three generative LLMs (Claude-3.5, GPT-4o, and LLaMA3.3-70B) demonstrated improved performance (sensitivity≥0.80, PPV≥0.82, and F1-score≥0.81) in identifying PTBM recommendations on the historical cohort test set (n=127) compared with the best performing pre-trained language model from our prior study (BioClinicalBERT),^15^ which achieved sensitivity=0.72, PPV=0.81, and F1-score=0.76 on the same test set. Claude-3.5 demonstrated the most balanced performance (sensitivity=0.86, PPV=0.86). GPT-4o achieved the highest PPV (0.91) with sensitivity=0.83. LLaMA3.3-70B had the lowest sensitivity (0.80) and PPV (0.82) (**Table 2**).

**Table 2.** Performance Evaluation on 2015-2019 Historical Cohort Test Set (n=127) and 2020-2024 Validation Cohort Test Set (n=122)

|  | Historical Cohort (2015-2019) |  |  | Validation Cohort (2020-2024) |  |  |
| --- | --- | --- | --- | --- | --- | --- |
|  | Claude-3.5 | GPT-4o | LLaMA3.3-70B | Claude-3.5 | GPT-4o | LLaMA3.3-70B |
| Sensitivity | 0.86 | 0.83 | 0.80 | 0.89 | 0.82 | 0.91 |
| Positive Predictive Value (PPV) | 0.86 | 0.91 | 0.82 | 0.95 | 0.97 | 0.89 |
| F1-Score | 0.86 | 0.87 | 0.81 | 0.92 | 0.89 | 0.90 |

### LLM Quantitative Evaluation: Validation Cohort (2020-2024)

Using the same selected prompt, all three LLMs demonstrated high performance in identifying documentation of PTBM recommendations in the validation cohort test set (n=122). Claude-3.5 achieved sensitivity=0.89, PPV=0.95, and F1-score=0.92. LLaMA3.3-70B had the highest sensitivity (0.91) with PPV=0.89 and F1-score=0.90. GPT-4o demonstrated the highest PPV (0.97) with lower sensitivity (0.82) and F1-score=0.89 (**Table 2**).

### Qualitative Evaluation of Model Explanations

Overall, the three LLMs excelled in comprehensiveness, agreement, and fabrication/falsification, and scored moderately on relevance. Performance varied for reasoning and logical sophistication, with Claude-3.5 demonstrating strong performance in both, while LLaMA3.3-70B and GPT-4o demonstrated lower performance, particularly for logical sophistication. **Table 3** presents examples of LLM-generated explanations rated by annotators as high, moderate, and low in overall trust, along with annotator justifications for each rating.

**Table 3:**
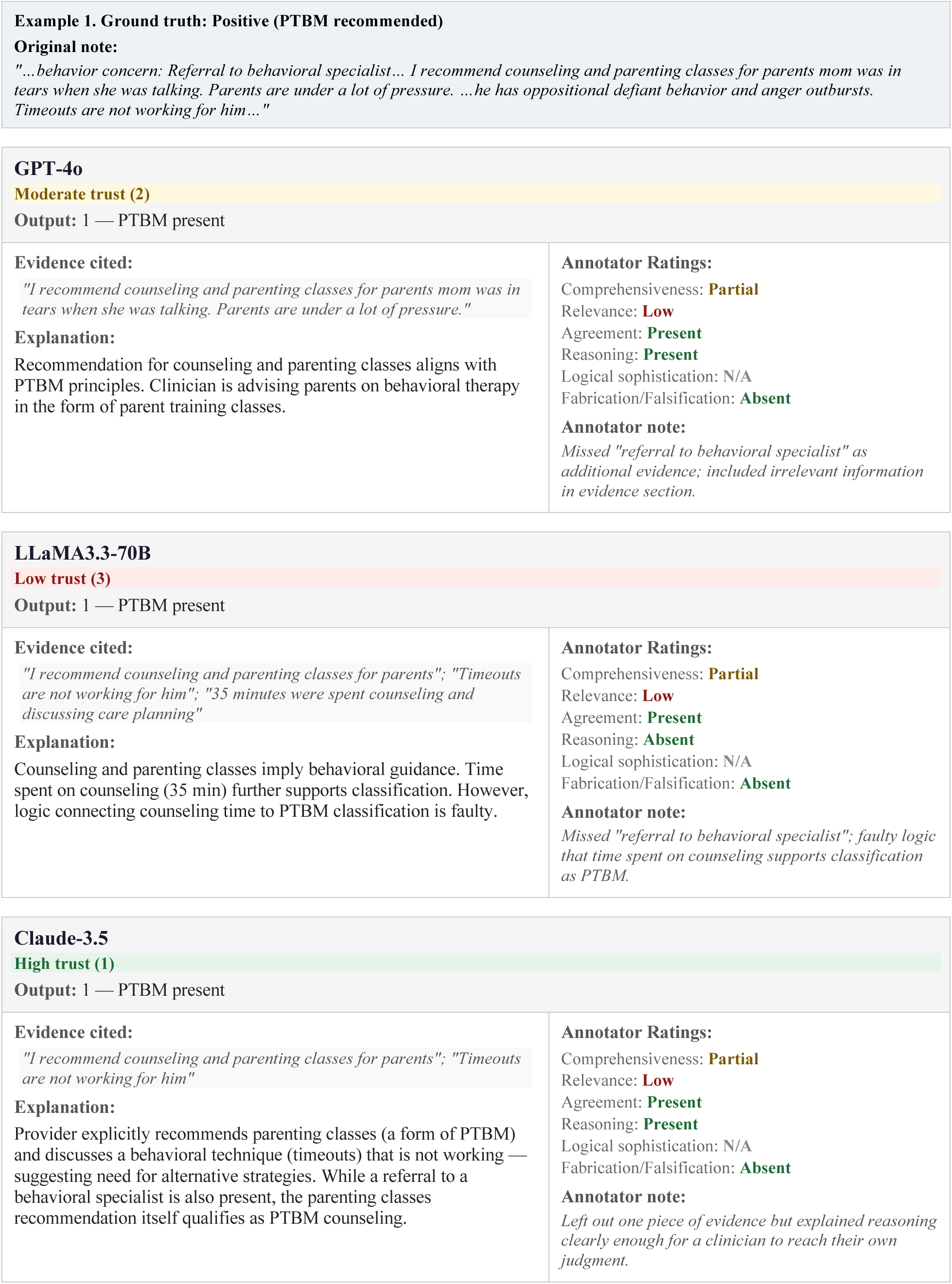
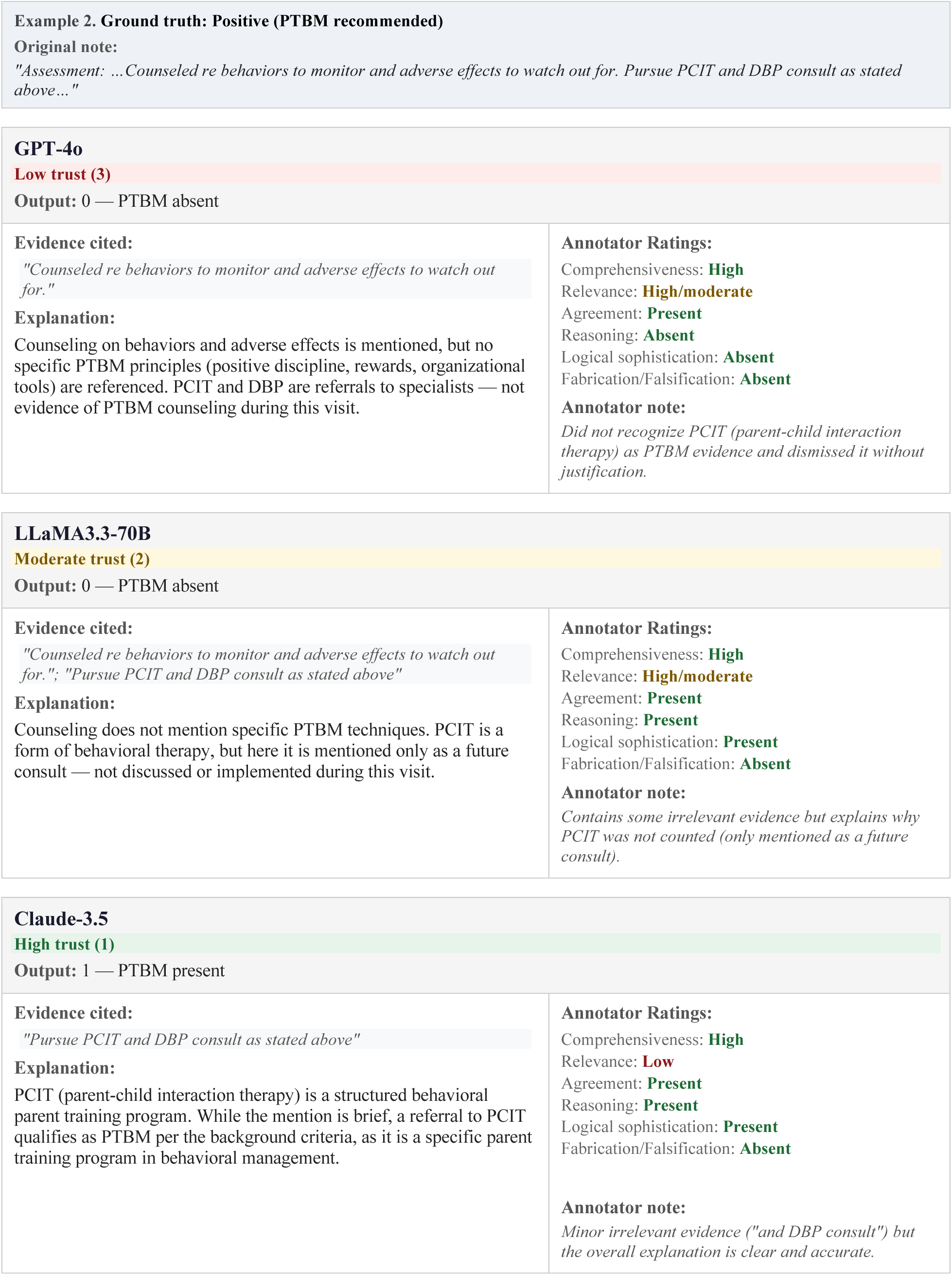
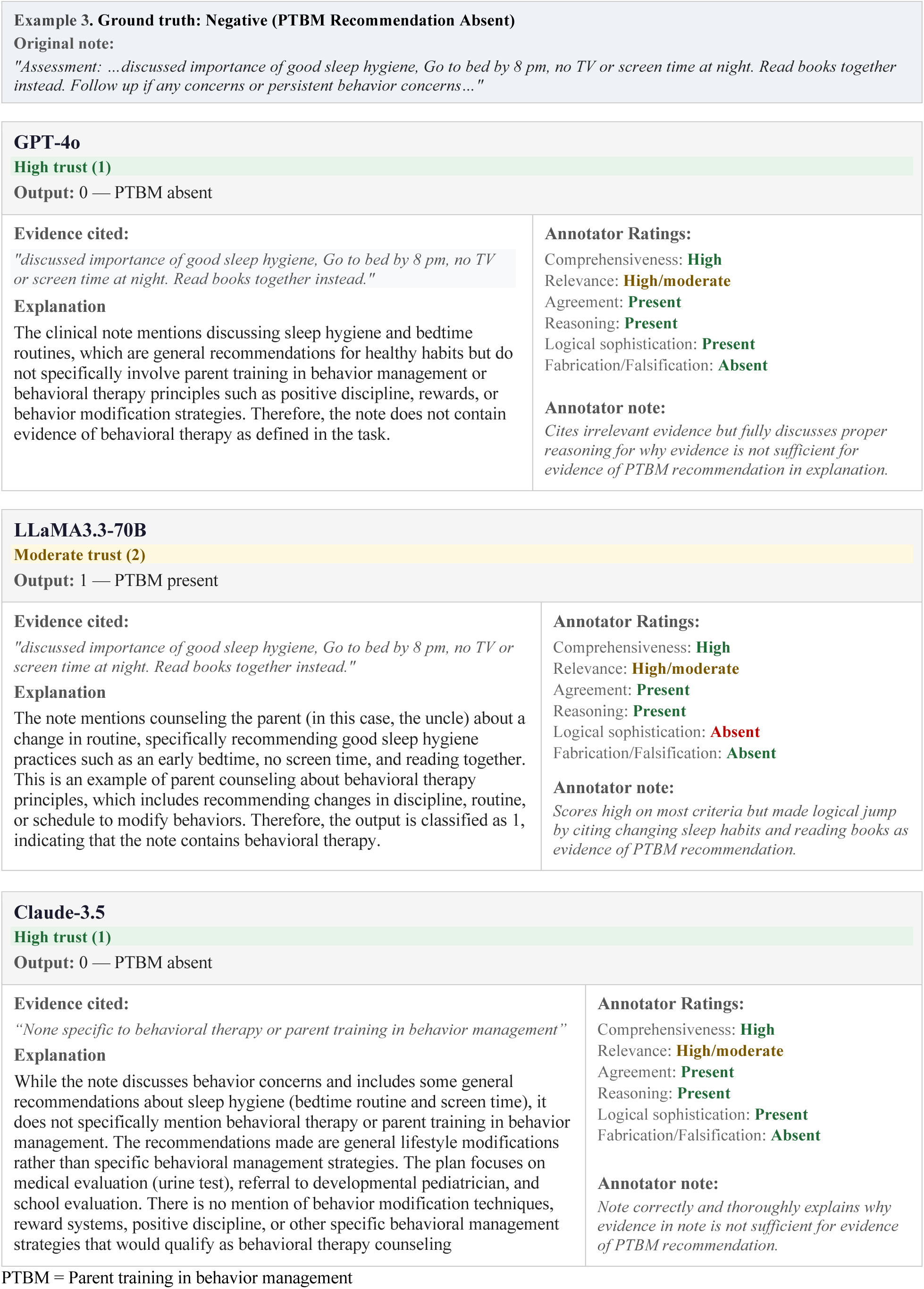
Examples of High-Ranking, Moderate-Ranking, and Low-Ranking LLM-Generated Explanations.

#### Comprehensiveness

All three models performed comparably well on comprehensiveness, citing relevant evidence for positive cases or justifying the lack of evidence for negative cases. For positive cases, GPT-4o scored highest (0.96), Claude-3.5 was intermediate (0.95), and LLaMA3.3-70B scored lowest (0.94); for negative cases, all models scored 1.

#### Relevance

Models demonstrated overall moderate performance on relevance, often citing evidence that did not match the gold-standard annotator evidence. For positive cases, GPT-4o scored highest (0.71), followed by Claude-3.5 (0.68), and LLaMA3.3-70B (0.63); for negative cases, all models scored 1.

#### Agreement

All models achieved excellent performance on agreement. For both positive and negative cases, all models scored 1, indicating full agreement within explanations with no contradicting statements.

#### Reasoning

Model reasoning ranged from moderate to excellent. Flawed reasoning included citing documentation of PTBM recommendation to justify negative classifications or relying on weak evidence to infer a positive classification. For positive cases, Claude-3.5 scored highest (0.97), followed by GPT-4o (0.96), and LLaMA3.3-70B (0.83); for negative cases, Claude-3.5 scored highest (0.81), then LLaMA3.3-70B (0.79), and GPT-4o scored lowest (0.61).

#### Logical Sophistication

Performance on logical sophistication varied widely. In many positive and negative cases, Claude-3.5 produced the most detailed explanations, integrating multiple pieces of evidence, recognizing nuances in phrasing, and synthesizing complex information. For positive cases, Claude-3.5 scored very high (1.0), LLaMA3.3-70B intermediate (0.70) and GPT-4o relatively low (0.67). For negative cases, Claude-3.5 scored high (0.87), LLaMA3.3-70B intermediate (0.83), and GPT-4o lowest (0.6).

#### Fabrication/Falsification

No fabricated or falsified statements were observed in all assessed notes, and all models scored 1 in this category.

#### Overall Trust

Annotators ranked Claude-3.5 highest for overall trust, noting the model consistently provided strong reasoning and cited relevant evidence from clinical notes and prompts (0.89). LLaMA3.3-70B had an intermediate ranking of overall trust (0.74), and GPT-4o ranked lowest (0.61).

#### Annotator Reflections on Ground-Truth Labels

Annotators also observed that, in a small number of cases, model rationales were sufficiently strong to prompt reconsideration of the original ground-truth label. These observations were anecdotal and not systematically evaluated, and no ground-truth labels were modified.

## DISCUSSION

In this retrospective cohort study, we demonstrated that large language models (LLMs) can accurately and transparently identify clinical recommendations documented in unstructured electronic health record (EHR) notes for children with ADHD. All three examined models demonstrated strong performance in identifying parent training in behavior management (PTBM) recommendations across two temporally distinct cohorts. Claude-3.5 achieved the most balanced performance in terms of sensitivity, positive predictive value, and F1-score, while also receiving the highest explainability ratings by annotators. This study, which requires replication in other health systems, demonstrates the potential of generative LLMs to offer scalable and continuous quality-of-care assessment, as an efficient alternative to manual chart review. Leveraging the demonstrated capabilities of LLMs may enable clinicians and health systems to receive transparent and near real-time feedback, which can facilitate targeted quality improvement efforts and mitigate persistent quality gaps in ADHD care.

The comprehensive evaluation of model explainability, guided by the QUEST framework, found that all examined models generated explanations that were comprehensive and relevant, demonstrated high agreement, and were free of fabricated or falsified content. While model hallucinations continue to present an important risk that requires human oversight in clinical applications of LLMs, narrowly scoped classification tasks and explicit prompting methods as used in this study may help reduce this risk that has been widely reported in prior studies.^22,23^

In other dimensions of explainability, particularly reasoning and logical sophistication, we found significant variation across models. These differences were most apparent in complex or nuanced clinical notes that required synthesis of multiple pieces of evidence or interpreting subtle context cues. Importantly, these distinctions were not always reflected in classification performance metrics, underscoring that models with similar accuracy can differ substantially in the coherence and depth of their explanations. This finding has important implications for LLM implementation in clinical and research settings, suggesting that relying on predictive performance alone is not sufficient. Prioritizing LLMs with strong explainability for implementation may contribute to clinician trust and adoption. Therefore, multidimensional evaluation frameworks that jointly assess accuracy and explainability will be essential for model selection and deployment in clinical settings.

Annotators observed that in some cases, model-generated rationales prompted the annotators to consider refining the original prompt and reconsider the original ground-truth label. This finding highlights the inherent ambiguity of some classification tasks that rely on available documentation. This finding also informs future implementation of LLM-based systems, highlighting the importance of identifying ambiguous documentation and maintaining human-AI workflows that support iterative task refinement based on clinician and LLM input.

While this study focused on a behavioral health application of LLMs, it offers a method that may be widely relevant for AI applications across clinical domains.^24,25^ The proposed method for transparent chart review addresses concerns regarding limited explainability of AI outputs that has been a major barrier to clinical adoption.^26,27^ The ability of LLMs to provide explicit rationales alongside classifications, established in the current study, may facilitate error and risk control by clinicians who can perform quality assurance spot checks. Future research is needed to evaluate whether implementing explainable AI workflows increases adoption and trust in AI-powered quality assessment.

### Strengths and Limitations

Strengths of this study include the evaluation of multiple modern LLMs using a single prompt, which facilitates a model-agnostic approach for clinical implementation and ongoing maintenance of rapidly evolving LLMs, and enables identification of ambiguous cases to be flagged for human review. The integration of both quantitative performance metrics and qualitative assessments of model explainability using a predefined framework is another study strength. Finally, the study’s focus on identifying quality gaps in pediatric ADHD care using real-world clinical documentation highlights the practical applicability of LLMs for scalable quality measurement in behavioral health – where patient care and outcomes are often documented only as free text.

This study has several limitations. The study was conducted within a single healthcare system, and documentation practices may differ across institutions, potentially limiting generalizability. Performance assessment was based on a sampling strategy enriched for model disagreements, which may not fully reflect performance in an unselected clinical population. Quality-of-care assessment relied on available clinician documentation of treatment recommendations, which may reflect documentation gaps rather than a lack of guideline adherence, potentially leading to misclassification. Despite the use of structured frameworks and consensus procedures for qualitative assessments of model explanations, ratings by trained annotators are inherently subjective, and assessment of the utility and acceptability of model explanations by clinicians is needed prior to implementation. Lastly, this study focused on a single point in time in a patient’s medical record (i.e., a single visit). Future work is needed to evaluate LLM performance in capturing longitudinal documentation across multiple visits, representing a patient’s trajectory of treatments and outcomes.

## CONCLUSIONS

LLMs can accurately identify treatment recommendations for young children with ADHD documented by PCPs in clinical notes, and provide reliable accompanying explanations, offering a scalable alternative to manual chart review for transparent quality-of-care assessment. This approach may contribute to overcoming longstanding barriers that have limited implementation of AI systems in pediatric clinical care.

## Supporting information

eSupplement

## Data Availability

The datasets generated and analyzed in the current study contain protected patient health information and are therefore not publicly available.

## ARTICLE INFORMATION

### Author Contributions

Drs. Bannett and Pillai had full access to all the data in the study and take responsibility for the integrity of the data and the accuracy of the data analysis.

*Concept and design:* Bannett. Pillai.

*Acquisition, analysis, or interpretation of data:* All authors.

*Drafting of the manuscript:* Pillai, Huang, Bannett.

*Critical review of the manuscript for important intellectual content:* All authors.

*Statistical analysis:* Pillai, Huang, Luo, Gunturkun.

*Obtained funding:* Bannett.

*Administrative, technical, or material support:* Pillai, Huang.

*Supervision:* Bannett.

## REFERENCES

1. Li Y, Yan X, Li Q, et al. Prevalence and Trends in Diagnosed ADHD Among US Children and Adolescents, 2017-2022. JAMA Netw Open. Oct 2 2023;6(10):e2336872. doi:10.1001/jamanetworkopen.2023.36872

2. Albert M, Rui P, Ashman JJ. Physician Office Visits for Attention-deficit/Hyperactivity Disorder in Children and Adolescents Aged 4-17 Years: United States, 2012-2013. NCHS data brief. Jan 2017;(269):1–8.

3. Danielson ML, Visser SN, Gleason MM, et al. A National Profile of Attention-Deficit Hyperactivity Disorder Diagnosis and Treatment Among US Children Aged 2 to 5 Years. J Dev Behav Pediatr. Sep 2017;38(7):455–464. doi:10.1097/dbp.0000000000000477

4. Wolraich ML, Hagan JF, Jr., Allan C, et al. Clinical Practice Guideline for the Diagnosis, Evaluation, and Treatment of Attention-Deficit/Hyperactivity Disorder in Children and Adolescents. Pediatrics. Oct 2019;144(4)doi:10.1542/peds.2019-2528

5. Charach A, Carson P, Fox S, et al. Interventions for preschool children at high risk for ADHD: a comparative effectiveness review. Pediatrics. May 2013;131(5):e1584–604. doi:10.1542/peds.2012-0974

6. Bannett Y, Gardner RM, Posada J, et al. Rate of Pediatrician Recommendations for Behavioral Treatment for Preschoolers With Attention-Deficit/Hyperactivity Disorder Diagnosis or Related Symptoms. JAMA pediatrics. 2021;doi:10.1001/jamapediatrics.2021.4093

7. Mittal S, Bax A, Blum NJ, et al. Receipt of Behavioral Therapy in Preschool-Age Children with ADHD and Coexisting Conditions: A DBPNet Study. J Dev Behav Pediatr. Dec 1 2023;44(9):e651–e656. doi:10.1097/dbp.0000000000001216

8. Bannett Y, Luo I, Azuero-Dajud R, et al. ADHD Diagnosis and Timing of Medication Initiation Among Children Aged 3 to 5 Years. JAMA Netw Open. Aug 1 2025;8(8):e2529610. doi:10.1001/jamanetworkopen.2025.29610

9. Shaw M, Hodgkins P, Caci H, et al. A systematic review and analysis of long-term outcomes in attention deficit hyperactivity disorder: effects of treatment and non-treatment. BMC medicine. Sep 4 2012;10:99. doi:10.1186/1741-7015-10-99

10. Alzu’bi AA, Watzlaf VJM, Sheridan P. Electronic Health Record (EHR) Abstraction. Perspect Health Inf Manag. Spring 2021;18(Spring):1g.

11. Casalino LP, Gans D, Weber R, et al. US Physician Practices Spend More Than $15.4 Billion Annually To Report Quality Measures. Health Aff (Millwood). Mar 2016;35(3):401–6. doi:10.1377/hlthaff.2015.1258

12. Schuster MA, Onorato SE, Meltzer DO. Measuring the Cost of Quality Measurement: A Missing Link in Quality Strategy. Jama. Oct 3 2017;318(13):1219–1220. doi:10.1001/jama.2017.11525

13. Tamang SR, Hernandez-Boussard T, Ross EG, et al. Enhanced Quality Measurement Event Detection: An Application to Physician Reporting. EGEMS (Wash DC). May 30 2017;5(1):5. doi:10.13063/2327-9214.1270

14. Hernandez-Boussard T, Blayney DW, Brooks JD. Leveraging Digital Data to Inform and Improve Quality Cancer Care. Cancer Epidemiol Biomarkers Prev. Apr 2020;29(4):816–822. doi:10.1158/1055-9965.Epi-19-0873

15. Pillai M, Posada J, Gardner RM, et al. Measuring quality-of-care in treatment of young children with attention-deficit/hyperactivity disorder using pre-trained language models. Journal of the American Medical Informatics Association : JAMIA. Apr 3 2024;31(4):949–957. doi:10.1093/jamia/ocae001

16. Feizollah A, Lin CY, O’Malley L, et al. The Use of Natural Language Processing to Interpret Unstructured Patient Feedback on Health Services: Scoping Review. J Med Internet Res. Aug 14 2025;27:e72853. doi:10.2196/72853

17. Shamszare H, Choudhury A. Clinicians’ Perceptions of Artificial Intelligence: Focus on Workload, Risk, Trust, Clinical Decision Making, and Clinical Integration. Healthcare (Basel). Aug 16 2023;11(16)doi:10.3390/healthcare11162308

18. Amann J, Vetter D, Blomberg SN, et al. To explain or not to explain?-Artificial intelligence explainability in clinical decision support systems. PLOS Digit Health. Feb 2022;1(2):e0000016. doi:10.1371/journal.pdig.0000016

19. Nong P, Platt J. Patients’ Trust in Health Systems to Use Artificial Intelligence. JAMA Netw Open. Feb 3 2025;8(2):e2460628. doi:10.1001/jamanetworkopen.2024.60628

20. Tam TYC, Sivarajkumar S, Kapoor S, et al. A framework for human evaluation of large language models in healthcare derived from literature review. NPJ Digit Med. Sep 28 2024;7(1):258. doi:10.1038/s41746-024-01258-7

21. Gruschow SM, Yerys BE, Power TJ, et al. Validation of the Use of Electronic Health Records for Classification of ADHD Status. Journal of attention disorders. 2016:First Published October 18, 2016. 10.1177/1087054716672337

22. Omar M, Sorin V, Collins JD, et al. Multi-model assurance analysis showing large language models are highly vulnerable to adversarial hallucination attacks during clinical decision support. Communications Medicine. 2025/08/02 2025;5(1):330. doi:10.1038/s43856-025-01021-3

23. Huang T, Tse G, Pageler NM, et al. Large Language Models Using Clinical Text in Pediatrics: A Scoping Review. JAMA Network Open. 2026;9(3):e262443–e262443. doi:10.1001/jamanetworkopen.2026.2443

24. Ford E, Carroll JA, Smith HE, et al. Extracting information from the text of electronic medical records to improve case detection: a systematic review. Journal of the American Medical Informatics Association. 2016;23(5):1007–1015. doi:10.1093/jamia/ocv180

25. Liu L, Blake V, Barman M, et al. Using natural language processing to extract information from clinical text in electronic medical records for populating clinical registries: a systematic review. Journal of the American Medical Informatics Association. 2025;doi:10.1093/jamia/ocaf176

26. Hassan M, Kushniruk A, Borycki E. Barriers to and facilitators of artificial intelligence adoption in health care: scoping review. JMIR Human Factors. 2024;11:e48633.

27. Hassija V, Chamola V, Mahapatra A, et al. Interpreting Black-Box Models: A Review on Explainable Artificial Intelligence. Cognitive Computation. 2024/01/01 2024;16(1):45–74. doi:10.1007/s12559-023-10179-8

