## Supplementary material for "Evaluating Large Language Models for Transparent Quality-of-Care Measurement in Children with ADHD": eSupplement

**eTable 1. Codes and Descriptors Used for Cohort Inclusion, Exclusion, and Defining ADHD-Related Visits**

| | ICD-10 Diagnosis Codes | ICD-10 Diagnosis Descriptors |
|---|---|---|
| **ADHD** | | |
| **Disorder-Level** | F90.0 | Attention-deficit hyperactivity disorder, predominantly inattentive type (and similar descriptions) |
| | F90.1 | Attention-deficit hyperactivity disorder, predominantly hyperactive type |
| | F90.2 | Attention-deficit hyperactivity disorder, combined type |
| | F90.8 | Attention-deficit hyperactivity disorder, other type |
| | F90.9 | Attention-deficit hyperactivity disorder, unspecified type (and similar descriptions) |
| **Symptom-Level** | F90.9 | Hyperactivity / Hyperactive behavior (and similar descriptions) |
| | R41.840 | Inattention (and similar descriptions) |
| | R41.844 | Executive function deficit |
| | R45.87 | Impulsiveness (and similar descriptions) |
| **ASD** | F84.0 F84.5 F84.8 F84.9 F94.8 F94.9 F80.82 | All |

ADHD = Attention-Deficit/Hyperactivity Disorder, ASD = Autism Spectrum Disorder

### eTable 2. Detailed Zero-Shot LLM Prompt

| **Prompt** |
| --- |
| You are reviewing a clinical note from health records of children with attention deficit hyperactivity disorder (ADHD) and classifying mentions of behavioral therapy.<br><br>**Background Information**:<br>- Behavioral therapy includes parent training in behavioral management, also known as behavioral therapy, behavioral parent training, or parent management training (e.g., triple p, incredible years).<br>- Parents can either be referred to a behavioral specialist or psychologist who offers parent training in behavior management or they can be counseled about behavioral therapy principles by the clinician who is writing the clinical note.<br>- Principles of parent training in behavior management includes positive discipline (positive reinforcement), system for rewards and consequences to modify behaviors, organizational tools such as visual schedules.<br>- Behavioral therapy does not include school-based interventions (e.g., 504 plan, school accommodations, daily report card, IEP, behavioral intervention plan).<br>- Behavioral therapy does not include psychotherapy such as cognitive behavioral therapy (CBT) or play therapy that do not include parent training or do not involve parent counseling about managing behaviors or behavior modification.<br>- Here are some examples of counseling parents about behavioral therapy: recommended change in discipline, routine, or schedule, or recommended use of rewards, incentives or checklists, discussed behavior modification, behavioral modification, focusing strategies, gave a handout on behavior management was provided to the family, recommended a book about behavior management (e.g., 1,2,3, magic).<br><br>**Task**<br>Please classify notes as containing behavioral therapy (1) or not (0). If there is no mention, then the output should be 0.<br><br>For each classification, provide the evidence from the note along with the classification output.<br><br>Provide a response in the following JSON format:<br>{"Output": 0 or 1,<br>"Evidence": [insert evidence for PTBM from note]<br>}<br><br>Process the following clinical note: |

**eTable 3. Annotation Guidelines and Scoring Rubrics for LLM Explainability Ratings**

| **Qualitative Scoring Criterion** (Related Concepts) | **LLM Output Sections that Criterion Applies to** | **Definition** | **Method of Evaluation** | **Annotation Guidelines** |
|---|---|---|---|---|
| **Comprehensiveness** (Completeness, exhaustiveness, complexity, additional information) | Evidence, explanation | Completeness of response provided by the LLM; The response should cover all critical aspects of the user's query, offering a complete overview or detailed insights as needed | **Likert scale 1-3**:<br>1 = high comprehensiveness<br>2 = partial comprehensiveness<br>3 = poor comprehensiveness | **True Positive cases**:<br>1 (High comprehensiveness) - meets both criteria below:<br>-Encapsulates all relevant pieces of evidence for PTBM<br>-The explanation explains most or all the cited evidence<br>2 (Partial comprehensiveness) - meets both criteria below:<br>-Missing one or more pieces of relevant evidence for PTBM.<br>-Explains most or all the cited evidence<br>3 (Poor comprehensiveness) - meets at least one of the criteria below:<br>-Completely missing any relevant piece of evidence for PTBM recommendation.<br>-None of the evidence cited is explained<br><br>**False Positive cases**:<br>Comprehensiveness is defined by how thoroughly a model explains the evidence it cites, even if the evidence is wrong:<br>1 (High comprehensiveness): The explanation explains all the cited evidence<br>2 (Partial comprehensiveness): The explanation explains some of the cited evidence<br>3 (Poor comprehensiveness): None of the evidence cited is explained<br><br>**True Negative and False Negative cases**:<br>1 (High comprehensiveness): Explanation clearly addresses and justifies the absence of PTBM evidence with reasoning<br>2 (Partial comprehensiveness): Explanation acknowledges absence of PTBM but provides only a minimal or vague justification |

| | | | | |
|---|---|---|---|---|
| | | | | 3 (Poor comprehensiveness): Explanation fails to address or explain the absence of PTBM evidence altogether |
| **Relevance** (Appropriateness) | Evidence, explanation | Alignment of response provided by the LLM to the user's query; The response should address the user's query without providing unnecessary or unrelated information | **Binary scale (1/0):**<br>1 = high/moderate relevance<br>0 = low relevance | **True Positive cases**:<br>1 (High/Moderate Relevance) - Meets both criteria below:<br>Evidence list does not include any extra irrelevant pieces of evidence.<br>Explanation stays focused on PTBM-related content<br><br>0 (Low Relevance) - meets at least one of the following:<br>Evidence list includes any extra irrelevant pieces of evidence<br>Explanation drifts into explaining things that are not PTBM-related without justifying their relevance<br><br>**False Positive cases**:<br>1 (High/Moderate Relevance): Even though the output and evidence are wrong, the LLM explanation stays focused on PTBM-relevant concepts (even if misinterpreted)<br>0 (Low Relevance): If explanation drifts into explaining things that aren't PTBM-related<br><br>**True Negative and False Negative cases**:<br>1 (High/Moderate Relevance) - meets at least one of the criteria below:<br>Evidence list is empty (e.g., "no evidence of PTBM"), or only notes ambiguous but potentially relevant content (and the explanation clarifies why it does not count)<br>Explanation stays focused on justification for why PTBM is absent without straying to irrelevant discussion (even if it missed relevant evidence)<br><br>0 (Low Relevance) - meets both criteria below:<br>Evidence list contains irrelevant content |

| | | | | |
|---|---|---|---|---|
| | | | | Explanation strays from explaining why PTBM is not mentioned in the note |
| **Agreement** (Alignment) | Classification, evidence, explanation | Coherence of response with established facts and theories; The response should be coherent and not contradict itself | **Binary scale (1/0):** 1 = agreement present 0 = agreement absent | **True Positive, False Positive, True Negative and False Negative cases:** 1 (Agreement present): No contradictions between the output, evidence, and explanation sections, AND no inconsistencies within each section 0 (Agreement absent): There was at least one contradiction between the output, evidence, and explanation sections OR within a section (evidence contradicts itself). |
| **Reasoning** (Logical coherence**,** logical error) | Explanation | Capability of the LLM to apply logical processing by linking the evidence to the output through clear reasoning steps | **Binary scale (1/0):** 1 = reasoning present 0 = reasoning absent | **True Positive and False Positive cases**: 1 (Reasoning present): Interprets the cited evidence in a logical and structured way with no logical errors and explains why the evidence supports a recommendation for PTBM. 0 (Reasoning absent): LLM does not show proper reasoning or logical processing steps, including only restating evidence with no or incorrect logic, providing vague or disconnected commentary, or jumping to the conclusion without explaining the connection.<br><br>**True Negative and False Negative cases**: 1 (Reasoning present): Explanation clearly describes the absence of relevant PTBM content and why that supports the negative classification. 0 (Reasoning absent): LLM gives a vague or unrelated reason for the negative classification or makes a classification conclusion without logic or supporting logical processing steps. |
| **Logical Sophistication** (nuanced explanation, interpretive sophistication) | Explanation | The ability of the LLM to synthesize and interpret multiple pieces of evidence in a balanced, | **Binary scale (1/0):** 1 = logical sophistication present 0 = logical sophistication absent | **Not applicable (N/A) if:** The clinical note is clearly positive or negative (i.e., no ambiguity, no competing or overlapping interpretations). If |

| | | | | |
|---|---|---|---|---|
| | | context-aware manner, reflecting a sophisticated understanding rather than a simplistic or literal response. | Not applicable (N/A) = note is very straightforward, and logical sophistication is not expected. | positive, the evidence is straightforward.<br><br>**True Positive and False Positive cases**:<br>1 (Logical sophistication present) - meets at least one of the criteria below:<br>Interprets ambiguous or borderline content and does not just repeat it.<br>Synthesizes or reconciles multiple pieces of confusing or partially conflicting evidence.<br>Demonstrates awareness of nuance, such as distinguishing school-based interventions from parent behavioral training or recognizing why general parenting advice is not sufficient.<br><br>0 (Logical sophistication absent) - meets at least one of the criteria below:<br>Explanation restates evidence without interpretation, ignoring or not explaining ambiguous content that needs clarification.<br>No synthesis or reconciliation of multiple pieces of confusing or partially conflicting evidence, if such are present.<br>Does not demonstrate awareness of nuance when needed.<br><br>**True Negative and False Negative cases**:<br>1 (Logical sophistication present): Recognizes potentially ambiguous content and includes interpretation that supports the negative classification.<br>0 (Logical sophistication absent): Explanation is overly simplistic and shows no effort to interpret possibly confusing or ambiguous information in the note or concluding absence of PTBM with no interpretive effort. |
| **Fabrication/Falsification** (Hallucination, confabulation) | Explanation | (a) Fabrication: the response contains entirely made-up information or | **Binary scale (1/0):**<br>1 = fabrication/falsification absent | **True Positive, False Positive, True Negative and False Negative cases:**<br>1 (Fabrication/falsification present): LLM states something |

| | | | | |
|---|---|---|---|---|
| | | data and includes plausible but non-existent facts in response to a user's query.<br>(b) Falsification: the response contains distorted information and includes changing or omitting critical details of facts | 0 = fabrication/falsification present | blatantly false (unrelated to its classification decision since that can be wrong), made-up, or distorted in explanation; For example, citing nonexistent information from the note.<br><br>0 (Fabrication/falsification absent); LLM does not have any fabrication or falsification in the explanation |
| **Overall Trust**<br>(Expert response, assurance, reliability, overall explainability) | Classification, evidence, explanation | The evaluator's overall confidence in the LLM response as a reliable, accurate, fair, safe, and useful response for interpreting clinical documentation. This rating considers clarity and perceived usefulness, especially in uncertain or nuanced cases. In addition, there is transparency regarding the LLM's capabilities and limitations | **Likert scale 1-3:**<br>1 = high trust<br>2 = moderate trust<br>3 = low trust | **True Positive, False Positive, True Negative and False Negative cases:**<br>1 (High Trust): You would confidently use or build upon this response in a clinical setting, even if the classification is incorrect; The explanation is clear, well-reasoned, internally consistent, and shows strong understanding of the clinical domain. Even if upon human review, it is determined that the model misclassifies, the explanation shows strong logic and depth of reasoning and provides a justifiable rationale based on the available evidence<br>2 (Moderate Trust): The response is mostly reasonable but has some limitations that lower your confidence; The explanation may be partially incomplete, slightly unclear, or mildly inconsistent with the classification or evidence but the reasoning is still understandable and grounded in the note.<br>3 (Low Trust): You would not rely on this response without major revision; The explanation is flawed, internally contradictory, unhelpful, or misleading. The model either misinterprets the note, misses obvious evidence, provides a weak or confusing explanation, or shows signs of hallucination or overconfidence. |

PTBM = Parent Training in Behavior Management
